# Influence of countries adopted policies for COVID-19 reduction under the view of the airborne transmission framework

**DOI:** 10.1101/2020.05.20.20107763

**Authors:** Charles Roberto Telles

## Abstract

Daily new cases dataset since January 2020 were used to search for evidences of SARS-CoV-2 community transmission as the main nowadays cause of constant infection rates among countries. Despite traditional forms of transmission of this virus (droplets and aerosols in medical facilities), new evidence suggests aerosols forming patterns in the atmosphere as a main factor of community transmission outside medical spaces. Following these findings, this research focused on comparing some countries and the adopted policy used as preventive framework for virus community transmission. Countries social distancing policy aspect, of one to two meters of physical distance, was statistically analyzed from January to early May 2020, and countries were divided into those implementing only social physical distance and those implementing distancing with additional transmission isolation (with masks and city disinfection). Correlating countries social distancing policy adoption with other preventive measures such as social isolation and COVID-19 testing, a new indicator results, derived from SIR models and Weibull parameterization, show that only social physical distance measure could act as a factor for SARS-CoV-2 transmission with respect to atmosphere carrier potential. In this sense, the type of social distancing framework adopted by some countries without additional measures might represent a main model for the constant reproductive spread patterns of SARS-CoV-2 within the community transmission. Finally, the findings have important implications for the policy making to be adopted globally as well as individual-scale preventive methods.

## 1. Introduction

### 1.1. The nonlinear properties of SARS-CoV-2 transmission patterns

As the COVID-19 epidemic is continuously demonstrating a growing pattern of community transmission, even with effective social isolation (lockdown or voluntarily shelter in place) and COVID-19 testing policies, theoretical analysis was performed to track the main transmission pattern of the virus reproductive behavior under other unsuspected forms of transmission (atmospheric) within social distancing policy and its infectious disease modeling concerning methodological aspects in the geographical regions samples of Asia, South America, North America, Middle East, Africa and European countries.

Focusing at advanced phases of community transmission observed in some countries and given the emergency of detecting the human transmission spreading patterns of the COVID-19 pandemic since December, 2019 and first quarter of 2020 [1], due to the increasing numbers of new infections and deaths monitored by the World Health Organization (WHO) [1] and John Hopkins University, this research mainly focused on the nonlinear epidemic properties of the population biology interactions and the highly random forms of virus transmission associated with human social behavior and environmental conditions (aerosol in long-range transmission (airborne transmission)). The nonlinear scenario pointed in this research refers mainly to the unpredictability of epidemiologic framework of SIR (susceptible, infected, removed) stochastic models to track the possible rate of infection among population even with some policy measures implanted along countries. This limitation of predict future rates of contagion was noticed during the pandemic spreading and it suggested the use of the qualitative theory of differential equations considering for it the high order non autonomous models to infer theoretically and without numerical results, for this moment, which of the variables that influences virus propagation present a possible fixed point orientation, as well as, explain why the nonlinearity can be tracked by this approaches. The random patterns of virus reproduction assumes through the air framework of analysis other dimension of research, conducting the social behavior of individuals and the fluid dynamics behavior as two main factors that produce still not resolved mathematical equations to simulate the virus daily new cases growing aspect.

SARS-CoV-2 expresses different patterns of transmission among humans [2-4]. This feature is being investigated not only using clinical trials and data [2-7], statistical tools [2,4,6-8], and medical interviews with patients [2,3,5-7], but also through the mathematical point of view. Concerning mathematical predictions of SARS-CoV-2 reproductive patterns, the maximum possible rate of infection of the virus and human daily life [2,9,10] among countries are presenting community spreading with an increasing margin of probability and statistical unpredictable outcomes. One of the unpredictable patterns (Figure 1) of the virus in its spread within the period of February to April 2020 is visible in the numbers of new infections among countries over time where the input and output, which is the possible number of people infected from an initial number, resulting into a maximum and minimum margins of spread of the virus fluctuation, express unpredictably (non-linearity). This observation was initially and briefly modeled by the nonlinear method reported by Gu et al. [11].The number of confirmed cases is lower than the number of total cases. The main reason for this is limited testing.

**Figure 1.**
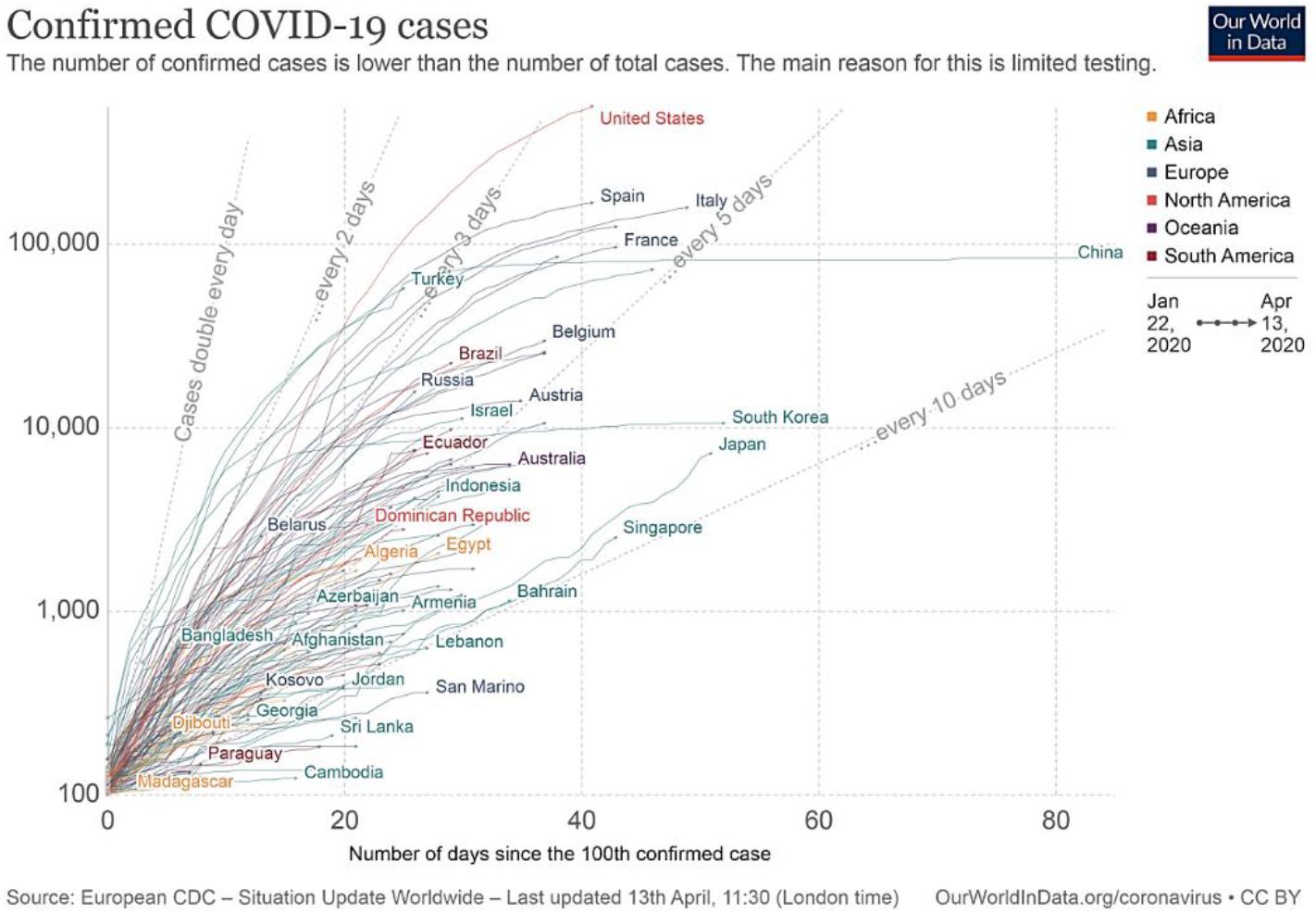
Total confirmed cases of COVID-19 worldwide: general overview. Observe the doubling of cases by day range and the large variance of input data for the evolution of each system. Source: Our World in Data.

Considering these nonlinear aspects of infection among countries, this article research during March and April [12,13,14] pointed to the airborne long-range transmission evidences of SARS-CoV-2 infection and this environmental role of infection was supported by the type of policies adopted in China and South Korea where significant reduction of infection cases occurred with distinct patterns found in other countries during all epidemics contagion phase. Regarding this two countries policies, social distancing measures with additional methods such as mask use and city disinfection were applied and this research found it as one main cause of COVID-19 non trivial frequency of daily new cases distribution during early stage of pandemics until end of April. In this sense, beyond the adoption of only social isolation and COVID-19 tests policies, social distancing with air preventive framework was revealed to be an urgent need for any country.

Furthermore, the variance observed in daily new cases among countries until the last date this research was performed, besides it is produced by reasons in each country, points of convergence (fixed point theoretical approach) are considered as stable concerning policies analysis point of view and with high stability towards many solutions obtained by the confounding variables as it will be demonstrated in Results section. These fixed point variables could be identified by the social isolation, COVID-19 testing and social distancing policies with atmospheric preventive framework. Even with high variance produced by other variables, these fixed point stable parameters can create a confident region of statistical analysis in terms of reducing maximum exponential growth of virus along days and therefore it could be more conclusive to many mathematical infectious disease models (SIR stochastic or deterministic approaches) that were created since the beginning of epidemics and later pandemics spreading.

The analysis of the nonlinear properties of the mathematical and non-pharmaceutical COVID-19 epidemiological framework is not only important for medical facilities but also for public policies and healthcare infrastructure to estimate the disease patterns of community transmission that are related to the potential damage within a pandemic scenario, which affects economics and threatens the health and survival of individual humans.

Finally, this research is relevant due to the large active workforce trying to maintain the minimum societal services and sectors necessary for survival, such as electrical, water, garbage, energy, food supply/production, commerce, and industry.

## 2. Methodology

### 2.1. Empirical evidence of instability for COVID-19 transmission: social isolation and social distancing differences

Recent studies reported that the transmission nature of SARS-CoV-2 is due to the proximity of other humans and social interactions within a set of empirical variables including the most basic forms of human behavior such as droplets in coughs, sneezes, handshakes, clothes, cups, general touching, and general objects-sharing behaviors [1,4,15,16]. This set of variables together with environmental factors of the virus’s possible transmission on the ground (surfaces) and in air (not only aerosol in medical facilities, but aerosol and biosols formation under atmospheric conditions for outside places) can also impacts transmission; therefore, generating new patterns for the course epidemiology [9]. This new pattern of virus transmission through aerosols was confirmed by the WHO in terms of a situation occurring at medical facilities [17] and also as stated by van Doremalen et al. [18], the upper and lower respiratory tracts in humans produce infection in the nearby atmosphere, propagating virus through the air, measured experimentally for about three hours during an experiment, and presenting low infection reduction at time of infectious titre from 10^3.5^ to 10^2.7^ TCID_50_ (tissue-culture-infective-dose) for SARS (Severe acute respiratory syndrome)-CoV-2. This overcoming and not predefined measure for an epidemic prevention demands alternative scientific hypotheses and further probabilistic and statistical frameworks needed to establish new policies and individual preventive actions.

This article new experimental research design provided an important approach to possible social distancing policies limitations due to no social transmission isolation of the virus, leading to the social distancing criteria of a physical distance between individuals with possible failures since air could lead to an unsuspected form of virus constant reproduction. The instability property could be visible when some countries in the adopted policy to contain virus spreading, fail to achieve this result, due to asymptotic instability between virus potential to infect individuals beyond this policy measures and methodology adopted. This feature has a wider domain of probability distributions assuming distinct patterns of occurrence [9,10,15] as observed in many countries [1], and also possibly contributing to the framework analysis of starting symptoms and final clinical consequences [4,6,7,19]. This observation was noted as the possible reason for the constant daily new cases and the community spreading of the disease that, even with preventive methods such as social isolation (lockdown or shelter in place) and COVID-19 testing, continued growing with exponential and unpredictable patterns are occurring.

The official pre-assumed forms of social distancing measures to avoid transmission and the possible new patterns of atmospheric disease factor may present a not yet observed continuous (not discretized) form of transmission (partially unpredictable) due airborne instability properties. This time-varying unresolved empirical data have high order behavior due to distinct nature of the variables for virus spreading and its feasibility at the moment, in terms of numerical results, is limited due to hard-to-calculate approaches involving human behavior, social behavior and virus forms of infection.

Therefore, other causes for virus daily new cases need to be considered beyond the traditional transmission analysis [2,6,10,20-22] if the rate of virus propagation continues to increase even with the implementation of social isolation and COVID-19 testing. Other unsuspected factors for transmission and modelling patterns should be considered and constructed [9-11,23-26] using mathematical counterproof predictions among countries that had already adopted social isolation, have COVID-19 testing available, but adopted social distancing measures with distinct parameters such as the use or not of masks with city disinfection. It is necessary to note that these three main policies are being executed worldwide as the main preventive measure to stop SARS-CoV-2 spreading [1].

The high chances of community transmission visible in the daily confirmed cases in statistical data random samples (table 1, figure 2) observed demands more empirical and theoretical investigations concerning how the virus gets transmitted and how hosts social behavior still contributes to the virus community spreading.

**Table 1.** Rolling three day average of daily new confirmed cases of COVID-19 among selected countries from 28-30 March, 11-13 April and 01-02 May, 2020. Source: Our World in Data.

| Countries | Average rolling three days new cases |  |  |
| --- | --- | --- | --- |
|  | 28-30 | 11-13 | 01-02 |
| United States | 19,011 | 32606↑ | 30399↓ |
| Spain | 7536 | 5054↓ | 1149↓ |
| Italy | 5717 | 4283↓ | 1974↓ |
| Germany | 5003 | 4092↓ | 1354↓ |
| France | 3673 | 3914↑ | 1116↓ |
| Iran | 2968 | 1814↓ | 1020↓ |
| United Kingdom | 2621 | 6086↑ | 5436↓ |
| Turkey | 1863 | 4647↑ | 2579↓ |
| Belgium | 1534 | 1538↑ | 566↓ |
| Switzerland | 1187 | 703↓ | 147↓ |
| Netherlands | 1145 | 1288↑ | 458↓ |
| Portugal | 806 | 948↑ | 343↓ |
| Canada | 746 | 1342↑ | 1682↑ |
| Austria | 595 | 279↓ | 72↓ |
| Brazil | 447 | 1600↑ | 6567↑ |
| Norway | 315 | 103↓ | 51↓ |
| Australia | 309 | 79↓ | 9↓ |
| Sweden | 298 | 577↑ | 633↑ |
| Denmark | 173 | 198↑ | 153↓ |
| China | 110 | 75↓ | 6↓ |
| South Korea | 110 | 29↓ | 6↓ |
| Finland | 87 | 139↑ | 103↓ |
| Singapore | 83 | 225↑ | 716↑ |
| Argentina | 77 | 114↑ | 135↑ |
| Chile | 255 | 460↑ | 881↑ |
| Saudi Arabia | 101 | 367↑ | 1340↑ |
| United Arab Emirates | 45 | 359↑ | 552↑ |
| Egypt | 31 | 126↑ | 284↑ |
| Pakistan | 117 | 238↑ | 1076↑ |
| Total Cases | 56,337 | 71619↑ | 60807↓ |

**Figure 2.**
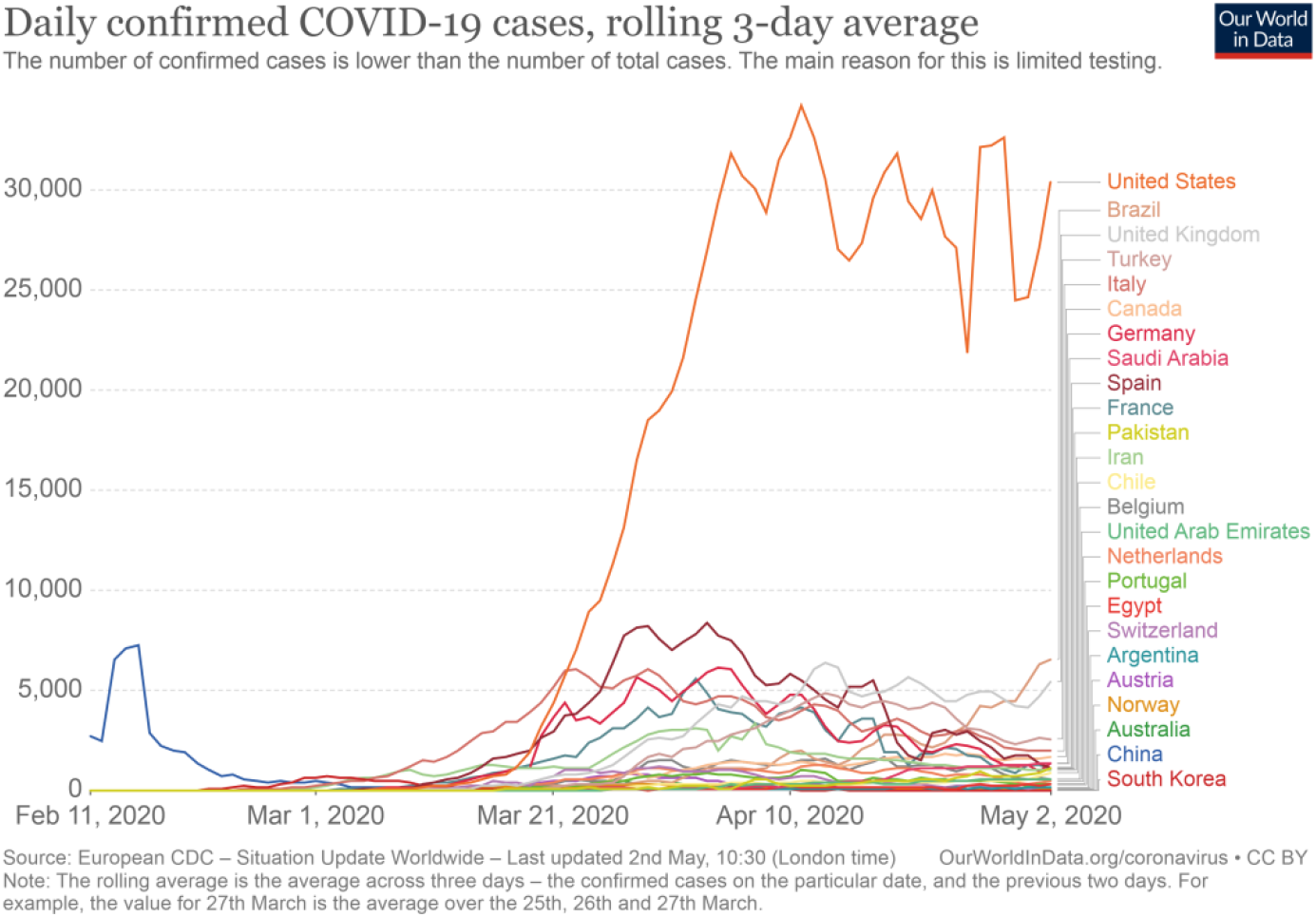
Table 1 statistical time series variance and average COVID-19 transmission cases among countries on 01-02 May, 2020. Source: Our World in Data.

### 2.2. Statistical uncertainty and the possibility of unified solutions to COVID-19 prevention

Many variables that affect virus transmission rates, such as the type of health policies adopted by each country, health infrastructure, population genetics, human variance in terms of biological resistance, epidemics outbreak, globalization aspects, COVID-19 testing availability, virus mutation, and citizen adherence to social isolation and social distancing present a strong influence on this figure 2 data. These confounding outcomes in each country pose a challenge to identify why some countries still have virus propagation active and what would be the best fixed point orientation to conduct all solutions to a desirable domain of virus contagion reduction. However these inputs of data can create a confounding environment of study, statistical worldwide data can provide a relevant confidence interval analysis if countries policies be compared, and thus revealing the best approach necessary to reduce virus infection. In this sense, it is justifiable the analysis policies adopted by countries as the most reliable, at the moment, form of reducing COVID-19 cases, while no vaccine or drugs present consistent and effective use for treating the disease or stop virus propagation.

For methodological reasons, many countries or regions, in the moment this research was performed, don’t present relevant data due to globalization aspects (i.e. some islands and some countries with low income per capita). Therefore they were not used for comparison since they did not present high infection daily cases and the urgency of policies measures to be adopted in large scale.

This research points of methodology indicates that individual behavior and social ties [27-29] are still the main focus of controlling the community spread of the virus, in respect to social isolation and social distancing measures, which consider the dynamics of groups/communities and community infrastructure (households, buses, shopping malls, meetings, markets, daily activities, and human behavior). Note that the term “social distancing” here is used for an uninfected individual outside medical facilities and refers only to the population separation patterns based on ground distances.

To explain this virus social transmission constant event, when social distancing is practiced, social contact might still occur as a human physical connection within environmental socialization, that is, physical ground and atmosphere contact. Most of the countries are adopting social distancing without the use of masks and city disinfection, aiming to restrict the social transmission of the virus. These policies expect that individuals distancing themselves in one or two meters, the desired effect of keeping people physically apart is enough to maintaining virus transmission ceased and also with the same effect as shelter in place (mandatory or not). With this measure, there are still many opportunities for social contact within a physical dimension regarding ground and atmospheric sense, both indoors or outdoors, as observed in many studies [16,30-34].

These two parameters, social distancing policy and social transmission isolation, were theoretically and empirically analyzed in this study because these variables are the possible forms of recurrent community transmission of the SARS-CoV-2. The epidemiological methods of prediction and control (which are the requirements for estimating the supply of financial, economic, and public health resources for the number of predicted infected people) lose their effectiveness due certain aspects of social transmission isolation and SARS-CoV-2 virulence potential [16,30-34]. This new approach diverges from other approaches as the one demonstrated by Hellewell et al [35], since social distancing and social transmission isolation parameters are considered in the approach, as well as the virulence under atmospheric conditions, which requires empirical results to be further investigated.

Many recent viral infectious diseases (SARS, MERS (Middle East respiratory syndrome), H1N1) share similar forms of transmission as SARS-CoV-2 [2], but they have different rates of exponential growth (Figure 3). Therefore, not only the causes of transmission, such as the chemical and biological properties of transmission and virus–human biological affinity issues (pharmaceutical interventions) must be considered, but the emergent virus and human social behavior events under environmental aspects [30-41]. And also, the nonlinear time series of worldwide policies can possibly present a clue for a high asymptotic stability (spreading network) [32] towards type of preventive policy measures adopted by each country as also observed previously with the *k* dispersion parameters and the superspreading prediction possibilities by Riou and Althaus [42].

**Figure 3.**
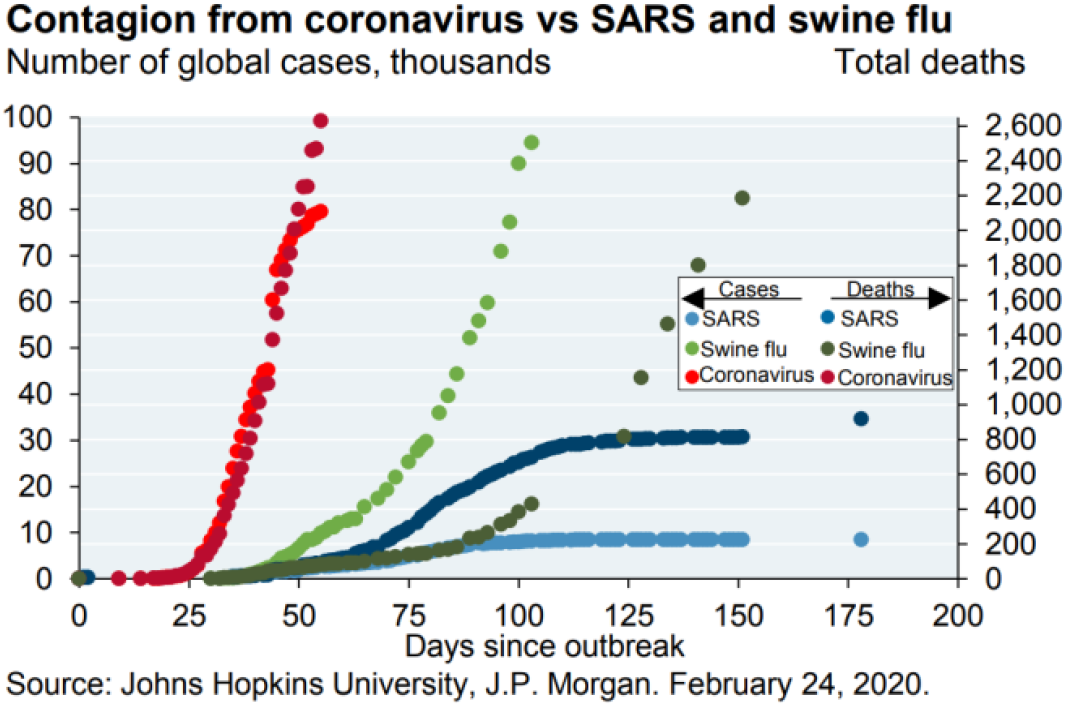
Viruses’ transmission behavior and rate of spreading patterns. Source: J.P. Morgan.

## 3. Results

These epidemiological factors (forms of transmission, biological-chemical affinities, and emergent social virus transmission behavior modeling patterns) ensures to the preventive epidemic framework [43] the need to consider any given number of infected individuals as potentially dangerous for pandemic initiation and continuation since uncertainty prevails. This led to the conclusion that there is no minimum range of infected individuals to consider the local epidemic as controlled; therefore, a post-critical epidemics event should be treated as an alert phase space, since new probabilistic outcomes are expected to generate the same recurrence plot of the non-autonomous phase space occurring in the epidemic origin (this research prediction was confirmed in many countries until early May [1]). Considering that epidemic scenarios have a natural upper limit and the posterior descendant tail has not yet been scientifically proven for this pandemic situation still as a natural ceasing of epidemic spreading, and therefore this theoretical observation should not be scientifically applied as a preventive measure.

### 3.1. Airborne transmission evidences

One main issue empirically observed in the media and also addressed in this research is related to the difference between the aspects of maintaining social distancing (e.g. physical distance at restaurants, parks, drugstore lines, household activities, house proximity (especially in slum neighborhoods), household tree proximity, markets, in house and outside social events through the windows and balconies, airplanes, ship balconies, hospital rooms, meetings, delivery or mail activities, prisons, residence, commerce, and industry in general) [44] and full social transmission isolation (with ground or atmospheric barriers). The daily media news and scientific reports [45,46] show that mostly China and South Korea and some other countries [45] have adopted mask use by citizens and full disinfection has been applied in crowded public spaces [11,47], and with some further concerns to public health professionals as reported by Li JP et al [48,49]. These policies actions are converging with the physical distancing criteria and possible failures, hence presenting physical transmission isolation barriers to airborne transmission (aerosol-biosols and atmospheric conditions [16,30-34]). At this point, a non-convergence effect of all these policies adopted by other countries would be when social distancing still present social activity at outside spaces without the use of masks and city disinfection. An overview about these non-convergence aspects can be observed in many researches. Concerning airplanes and community policy actions was provided by Chinazzi et al. [50]. In this sense, the social connection that occurs might be one of the unobservable factors of transmission if virus potential to spread under atmospheric conditions [30,31,51-54] is still active through air fluid [16,30-34] and not only ground preventive framework. Mostly of the recommendations for physical distance address the virus’s potential to spread through ground and air contexts based on human liquid droplets, but not air–fluid complex scenarios without droplets involved (e.g. pollution). Concerning recent atmospheric data and SARS-CoV-2 relationships, Wickramasinghe et al. [51] reported several cases of person-to-person transmission patterns in which in this research view can be understood by the lack of virus social transmission isolation policies in terms of additional barriers, such as masks and city disinfection, which led to air transmission. This observations were also pointed by Cembalest [52] atmosphere data and a brief analysis and Pirouz et al. [53] mathematical modelling, with a deep analysis of how the atmosphere parameters of temperature, humidity, and wind affect population density output for SARS-CoV-2 infection, guiding these studies to the proximal conclusion of the strong impact of atmosphere on the actual non-understood patterns of virus community constant dissemination among countries that do not use mass mask policies with city disinfection, but adopted social distancing criteria. A final research that leads towards this research results was Poirier et al [54] who examined the weather conditions capable of generating the full transmission patterns observed in Figures 1 and 2, and the lack of social transmission barrier for the airborne transmission variable.

### 3.1.1. Maximum exponential growth and duration of epidemics in days as evidence

This statistical data of Figure 4 show the rise of daily new cases around the world with all policies adopted by countries such as social isolation, COVID-19 testing, and social distancing criteria, associated or not with the use of masks and city disinfection.

**Figure 4.**
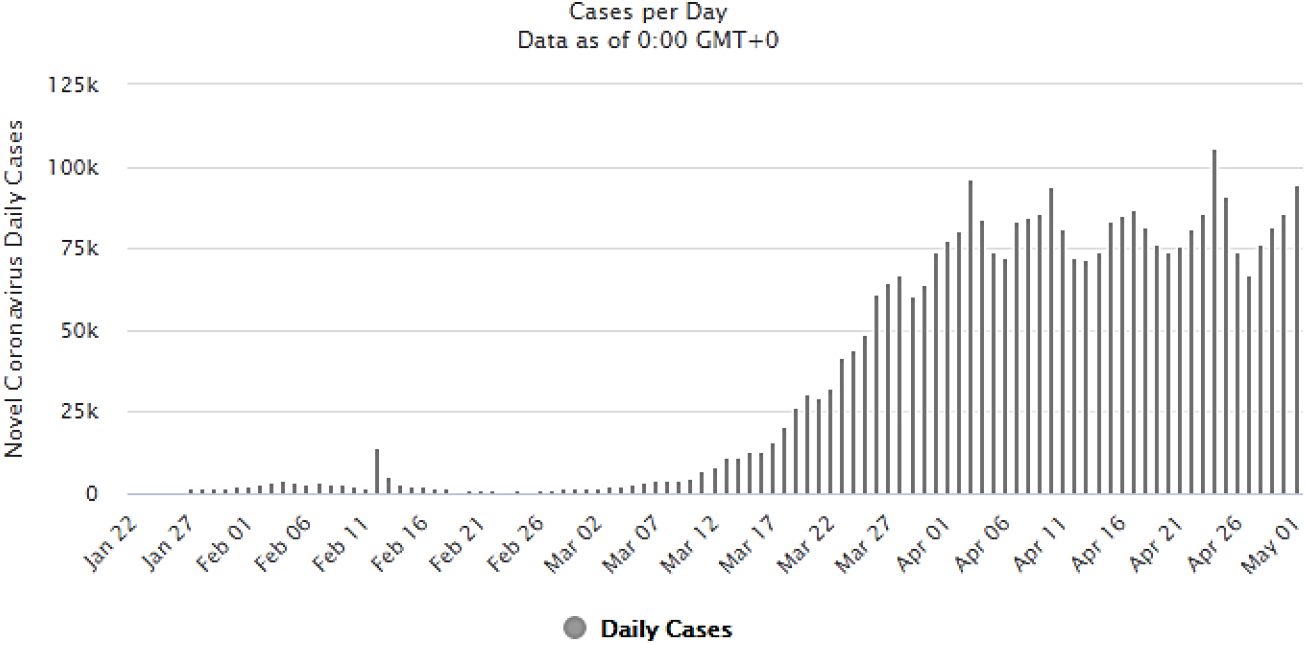
From February to May, 2020, general overview of all reported cases of COVID-19 worldwide. Source: Worldometer.

Besides many of European countries are adopting different measures for prevention or availability to it, one specific point beyond social isolation and COVID-19 testing can be highlighted. These countries, until the date this research was conducted, have not introduced mass mask use and or constant city disinfection as was adopted by China and South Korea from the early to later pandemic spread. This inference is supported also in other countries when on late March and during April [1], the infection rates in countries such as Italy, Spain, Iran, United States, Germany, France, and Brazil present constantly generating new daily infections with different patterns of China and South Korea, something that is still visible for these countries on May, 2020 so far.

In Figure 5 European countries, even with social isolation, COVID-19 testing availability, and social distancing measures, in late March and start of April, despite many citizens not obeying/adhering to institutional orders, reports indicated a reduced number of citizens outside homes but daily constant infections cases over 30.000 population during April (maximum exponential growth with growing patterns for a long period, for a total of 58 days from 28 February to 25 April, 2020.

**Figure 5.**
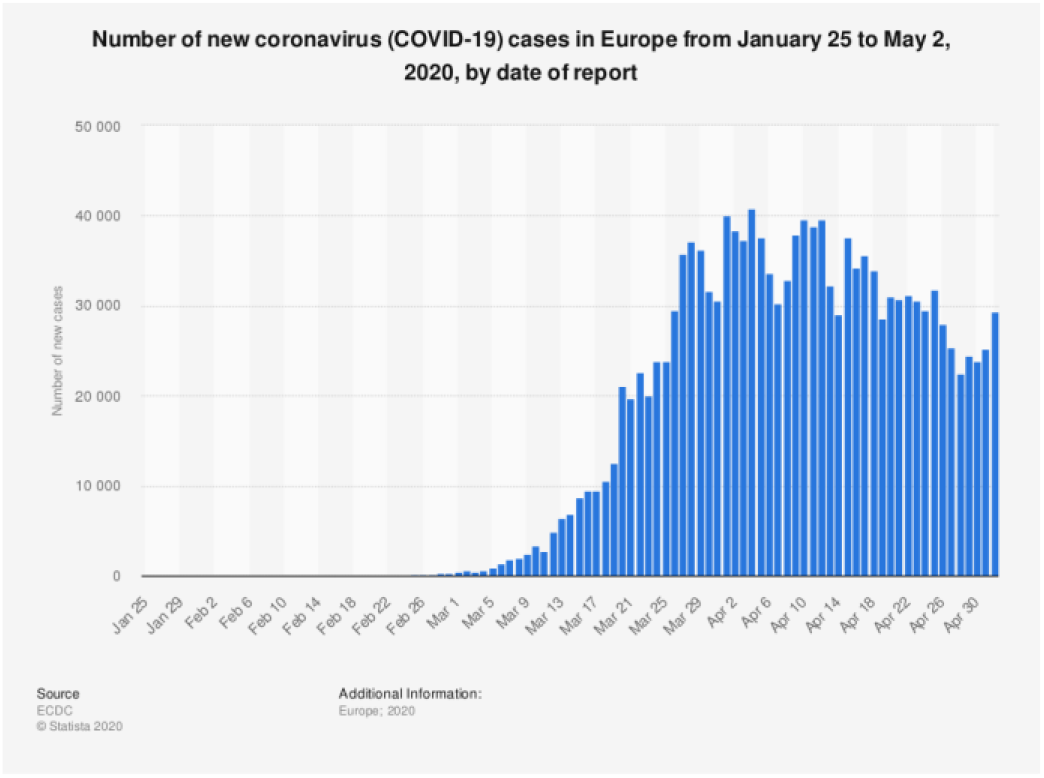
COVID-19 new infection cases day by day in Europe. Source: Statista.

Observing European cases in Figure 5 and in Italy specifically (Figure 6), concerning individuals that do not obey the direction or obligation to stay at home, these uncertain individual actions can generate several random transmission outputs as well. These specific random aspects contribute to the statistical variance of the countries, including in the number of infected and the mortality.

**Figure 6.**
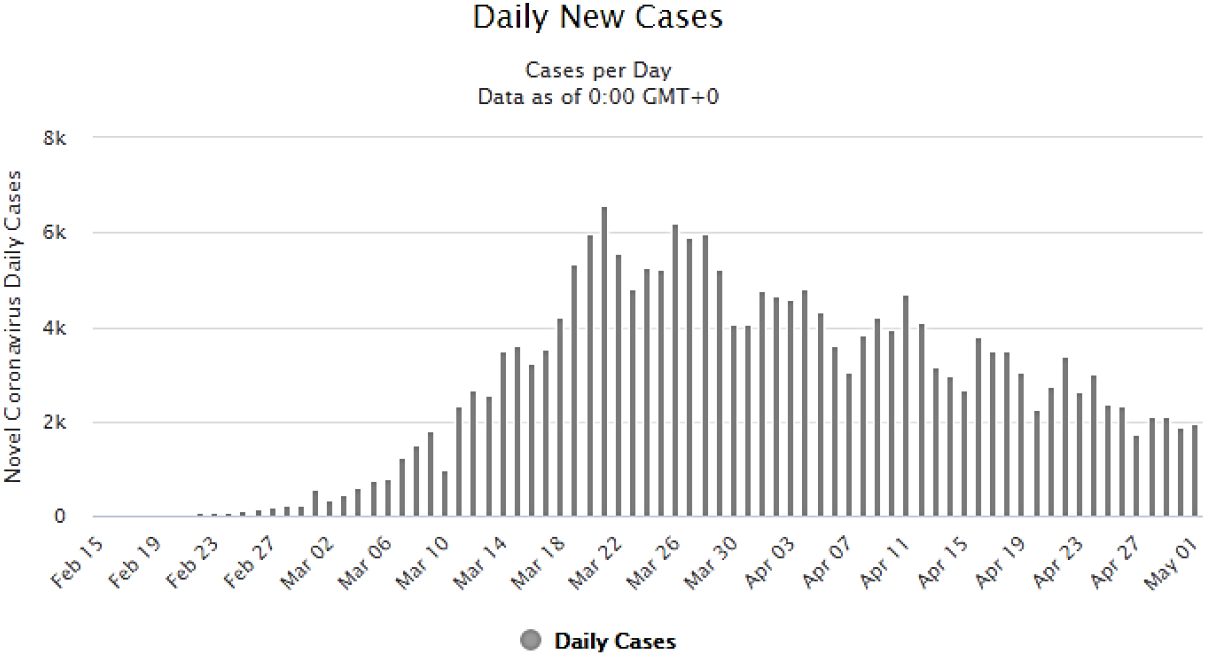
New daily COVID-19 cases in Italy. Note that Italy mask use policy for population was introduced by ending March and April start, being this measure carried out until the last date this research was conducted (05/02/2020). Source: Worldometer.

Analyzing figure 6, Italy counting days for the exponential growth, it express daily constant infections cases with growing patterns rates starting from the epidemic outbreak until 4.000 population (maximum exponential growth rate for a long period of 51 days from 22 February to 12 April, 2020.

Many other factors are being discussed nowadays to explain why those countries are still presenting a rise in the virus propagation, such as testing availability and the date the city started social isolation measure. Besides it can also have strong influence on the virus undetected phase of exponential growth, the time series of this statistical data can give also a glance about countries divergences for how much time was needed to each country to stabilize its virus infection. And this will be still the focus of analysis in the next days. Observing preventive methods concerning airborne vectors, which were adopted by South Korea (Figure 7) and China (Figure 8) as the default (masks and city disinfection) the virus social transmission behavior reproductive patterns for these areas differ.

**Figure 7.**
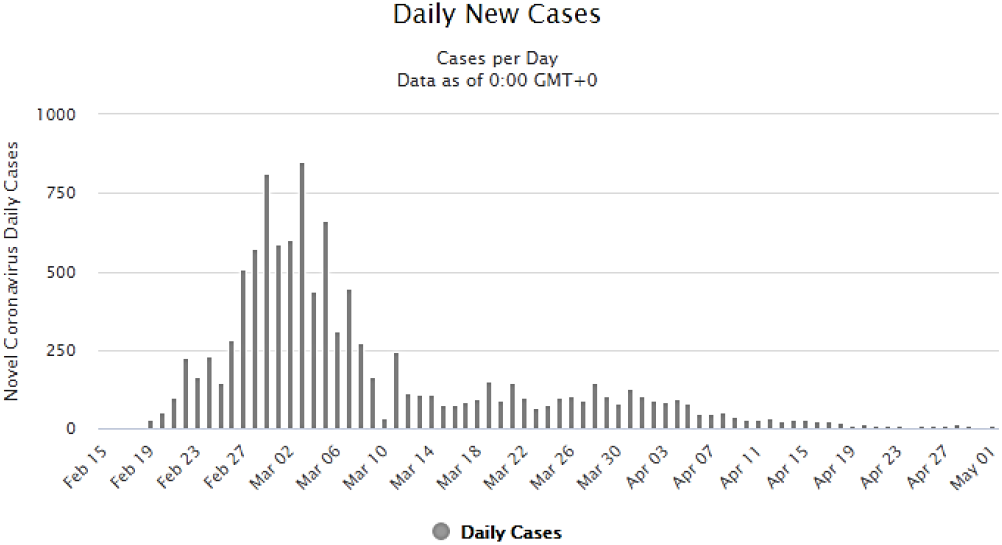
Daily new cases of COVID-19 in South Korea. Source: Worldometer.

**Figure 8.**
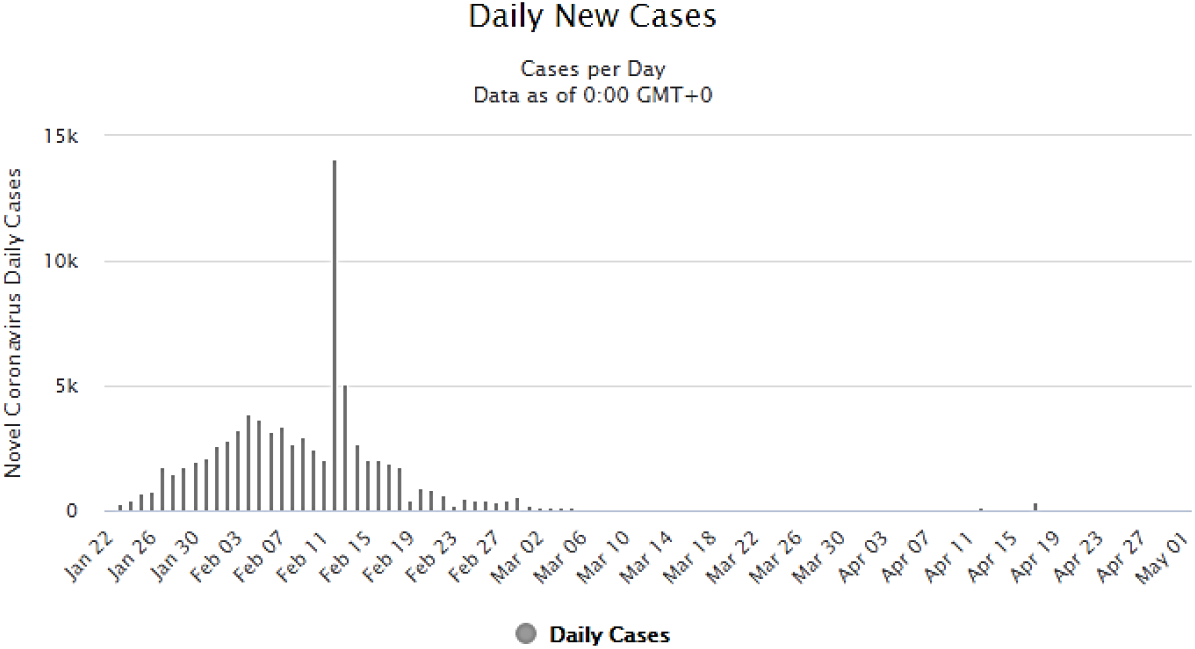
Daily new cases of COVID-19 in China. Source: Worldometer.

Figure 7 and 8, these countries adopted social distancing with air preventive measures, with a total of 20 days of maximum exponential growth rate over 250 population (19 February to 8 March, 2020) for South Korea and with a total of 28 days of maximum exponential growth rate over 1.500 population (23 January to 18 February, 2020) for China.

And also important to say it is necessary to consider exceptions for a possible micro-dimension of analysis of population biology that can occur in any country, as a local problem [55-57] that might sometimes count or not towards a high exponential growth rate of virus transmission. China experienced high exponential virus transmission growth due to the initial conditions of the new disease; country needed time to evaluate and adopt policies and scientific measures, as observed by Pan et al. [57].

#### 3.2.2. Maximum exponential growth mean and spreading rate over day’s evidence

An overview about these explained countries maximum exponential growing patterns and counting days are available at Table 2. data with other countries included. Table 2, with the same scheme of data extraction as presented in figures 5-8, for the first column presents the maximum growth of infection obtained by the maximum exponential mean reached in average days since the outbreak and therefore not counting for higher growths than the mean “y” that some countries present (more explanation will be given in third column details). The second column “ t” presents how much days the infection presented an exponential growth with a maximum mean reached. The third column data collection reflects the maximum exponential infection spreading rate over days with the following theoretical design involving SIR models and missing gaps of this models for this specific COVID-19 disease.

This third column approach have similarities with SIR models, but it is based on distinct aspects of analysis of variables S and R by removing these variables from the formula and focusing mainly in the I variable defined by Weibull parameterizations and exponential distributions. This design of analysis is very relevant due to instability aspects of SIR analysis taken since outbreak of disease, mainly existing in the S and R variables due infodemics, type of analysis conducted for this research (the apparent lack of overall topological homology of data) and other nonlinear aspects of the new coronavirus 2 disease. For this reason, the proposed method of analysis considers only the infectious disease aspect of evolution of cases rather than pre assuming full immunity or deterministic models for population behavior that is in this case one of the most influencing forms of keeping the virus active in its propagation.

**Table 2.** COVID-19 maximum exponential growing patterns/population and time period by country/region within 07 April to 01 May, 2020. Note that some countries are currently with their max. exponential infection spread active in the day research was conducted (different epidemic phases). For these cases, no final exponential score was reached yet (marked in yellow color), however this not count for future predictions. The *y* and *t* data showed in table are retrieved for 01 May. Data source: Worldometer.

| Country | $y$ | $t$ | $R$ on 7 April | $R$ on 13 April | $R$ on 01 May |
| --- | --- | --- | --- | --- | --- |
| Worldwide | 60.000 | 99 | 675,67 | 886,07↑ | 606,06↓ |
| Europe | 30.000 | 91 | 410,95 | 379,74↓ | 329,67↓ |
| Italy | 4.000 | 51 | 90,90 | 78,43↓ | 78,43= |
| South Korea | 250 | 20 | 12,5 | 12,5= | 12,5= |
| China | 1.500 | 28 | 53,57 | 53,57= | 53,57= |
| Iran | 2.000 | 46 | 44,44 | 43,47↓ | 43,47= |
| Spain | 5.000 | 55 | 131,57 | 121,95↓ | 90,90↓ |
| France | 4.000 | 45 | 138,88 | 100↓ | 88,88↓ |
| United States | 30.000 | 53 | 625 | 540,54↓ | 566,03↑ |
| Brazil | 5.000 | 51 | 25 | 28,57↑ | 98,03↑ |
| Germany | 4.000 | 41 | 105,26 | 100↓ | 97,56↓ |
| Russia | 5.000 | 46 | 15,62 | 27,02↑ | 108,69↑ |
| United Kingdom | 5.000 | 58 | 64,51 | 69,64↑ | 86,20↑ |
| Singapore | 500 | 57 | 0,96 | 1,75↑ | 8,77↑ |
| Portugal | 500 | 49 | 15,15 | 13,15↓ | 10,20↓ |
| India | 1.000 | 58 | 8,33 | 9,43↑ | 17,24↑ |
| Canada | 1.500 | 48 | 27,77 | 24,39↓ | 31,25↑ |
| **Japan** | 500 | 67 | 3,92 | 7,14↑ | 7,46↑ |
| **Sweden** | 500 | 58 | 5,71 | 0,65↓ | 8,62↑ |
| **Argentina** | 100 | 57 | 1,51 | 2,56↑ | 1,75↓ |
| **Chile** | 500 | 52 | 10 | 8,33↓ | 9,61↑ |
| **Saudi Arabia** | 1.000 | 54 | 3 | 4,16↑ | 18,51↑ |
| **United Arab Emirates** | 400 | 70 | 4,34 | 5,76↑ | 5,71↓ |
| **Egypt** | 200 | 62 | 1,31 | 2,27↑ | 3,22↑ |
| **Pakistan** | 500 | 53 | 3,44 | 5,71↑ | 9,43↑ |

For this analysis, it was considered the observed infected population samples *Y* = (*Y*_1_,…*Y_n_*) of the exponential growth *f(Y;λ*) = *λe*^−^*^λY^* where the samples are taken from zero cases until the observed maximum exponential mean reached for each country with unknown predictive scale of *exp λ* or maximum likelihood estimator for *λ* due to nonlinear outputs generated for *Y* with heteroskedasticity. In this simple form, defining the mean as *y =* 1/*λ*, the numerical representation of the ratio between days and the mean can be obtained by observed exponential mean scale until it reaches a form like having adopted for the calculations with conditional shape of Weibull parameterization like *κ* < 1. At this point the days counting forward this condition will be rejected to extract the maximum exponential infection spreading rate in the formula *rate* (*R*) = *y/t* and *t* = *κ* and only in the desired event expression. This approach can be more sensitive in terms of time overview of the disease and its potential to infect as time passes. This sensitivity is much more confident for predictions due to the exponential behavior of infections at community phases.

Note that at Table 2, some countries present lower exponential growing rate than China or South Korea. These data need to be observed in terms of country starting epidemic outbreak, country epidemic time period and mainly countries that are currently active its maximum exponential growth. For this later feature, some countries did not reached yet a valid data to measure if country policies already helped to flatten the curve of daily new cases and they can present active exponential raise, therefore, further future analysis is required to able to perform a confident comparison. China being the first country to elaborate countermeasures policies, might presented some delay and therefore maximum exponential rate was reached before these measures could take effect. Also, many countries that adopted known pre-established measures presented better performances than the world epidemic starting ones, but since they remain with active and low exponential growth, as an example, Singapore with low max. output of exponential rate, they did not reached the same results as China and South Korea with the adoption of additional preventive measure for social distancing/city disinfection and the high reduction of virus exponential spreading patterns. This Singapore scenario occurs in many other countries as well. Also Singapore presented a rise in the max. exponential growth from 50 (07 April) to 500 (02 May). Germany, Italy, Portugal, Iran and France presented until the day the data was retrieved an end in the mean maximum exponential rate reached, however this does not count for future epidemics behavior to be observed as a deterministic approach. Further analysis are required along pandemic worldwide.

By observing figures 5–8, it is possible to evaluate how long it took for some countries that implemented social distancing measures plus airborne preventive methods to flatten the exponential growth of community infections. Countries that only applied social distancing without mask use and city disinfection at early stages required many more days than other countries that applied different measures. Also, many other scenarios are observed since countries policies towards mask use and city disinfection are still on implementation phase.

Also one more important note on this table 2 data refers to different epidemic phases of data collection for each country. These distinct phases are important to be considered together due to need of a methodology that can extract behavior of the disease in the not optimal evolution (deterministic) of the virus infection and policies adopted by countries, hence, revealing in the complex scenario the disease dynamics under a confounding environment of data.

#### 3.2.3. Maximum exponential growth mean X time X cases per 1 million population

Table 3 as of 07 April to 01 May range, 2020 (Worldometer), shows the comparison of how many cases occurred in the selected countries of analysis and it is possible to observe China and South Korea low cases per one million population, low duration of epidemics and with stable exponential growth for both countries. Notably some countries present lower cases per one million population, but they are all with growing patterns of infection propagation, longer duration of epidemic and with high exponential growth rate pattern. In this point, it is possible as a third evidence, to state that China and South Korea reached with specific policies adoption until the moment this research was performed, the best score for the correlation between total cases per one million population with country time period of infection and COVID-19 growing patterns stability. Note that any range of analysis to be performed will have its values of time and max. exponential mean modified according to the selection taken. The higher the range, the better is *R* precision.

**Table 3.** Some countries with COVID-19 spreading and total infected cases per one million population. The *t* used is counted for 01 May, 2020. Data source: Worldometer on 01 May, 2020.

| Countries | Total Cases | Total Cases/1 M population | $t$ | $R$ from 07 April to 01 May |
| --- | --- | --- | --- | --- |
| USA | 1,159,430 | 3,503 | 53 | 566,03↑ |
| Italy | 209,328 | 3,462 | 51 | 78,43= |
| China | 82,875 | 58 | 28 | 53,57= |
| Spain | 245,567 | 5,252 | 55 | 90,90↓ |
| Germany | 164,967 | 1,969 | 41 | 97,56↓ |
| Iran | 96,448 | 1,148 | 46 | 43,47= |
| France | 168,396 | 2,580 | 45 | 88,88↓ |
| UK | 182,260 | 2,685 | 58 | 86,20↑ |
| Sweden | 22,082 | 2,186 | 58 | 8,62↑ |
| India | 39,699 | 29 | 58 | 17,24↑ |
| Japan | 14,305 | 113 | 67 | 7,46↑ |
| S. Korea | 10,780 | 210 | 20 | 12,5= |
| Russia | 124,054 | 850 | 46 | 108,69↑ |
| Singapore | 17,548 | 2,999 | 57 | 8,77↑ |
| Portugal | 25,190 | 2,470 | 49 | 10,20↓ |
| Canada | 56,714 | 1,503 | 48 | 31,25↑ |
| Brazil | 96,559 | 454 | 51 | 98,03↑ |
| Argentina | 4,532 | 100 | 57 | 1,75↓ |
| Chile | 18,435 | 964 | 52 | 9,61↑ |
| Saudi Arabia | 25,459 | 731 | 54 | 18,51↑ |
| United Arab Emirates | 13,599 | 1,375 | 70 | 5,71↓ |
| Egypt | 6,193 | 61 | 62 | 3,22↑ |
| Pakistan | 19,022 | 86 | 53 | 9,43↑ |

In table 3, orange lines indicates the best scores reached by two countries while yellow colors indicates countries that reached best score compared to China and South Korea. Observe that even with good scores for some countries, they do not fit for all table columns and also present exponential growing rate as retrieved data for 01, May, 2020. Despite many countries have close proximity to China and South Korea and they do not fit these later countries results, several factors influence the numbers oscillations and differences as described in methodology section. Notably, Argentina performed the best score of South America and many other regions worldwide. Voluntarily and later obligatory mask use and city disinfection leaded Argentina to the same epidemic scenario obtained by China and South Korea, reaching successful results. United Arab Emirates and Portugal, by presenting decreasing exponential growth rate, could reach better results with the introduction of air preventive measures.

Clearly table 3 present much of the unpredictability based on nonlinear factors such as health policies adopted by each country, health infrastructure, population genetics, COVID-19 testing availability, and citizen adherence to social isolation/social distancing. These data indicate further studies are necessary to obtain more accurate numerical results, since each country undergoes a period of disease spread with different rates as mentioned four paragraphs before. Despite these variances producing large differences in the outcomes, most of the countries adopted social distancing as a method of virus spread prevention with no obligation to social isolation, which became a default pattern for prevention in later February and early March, this also contributed to the virus incubation period and spreading patterns rates to increase much more than observed in China and South Korea. These results points to the conclusion that besides many factors influence in the outcomes, some specific patterns are divergent in these two countries compared to the others, since they present more lower duration in days of epidemic, stable (also low) disease exponential growth patterns and low confirmed cases per 1 million population as indicated in figure 9.

**Figure 9.**
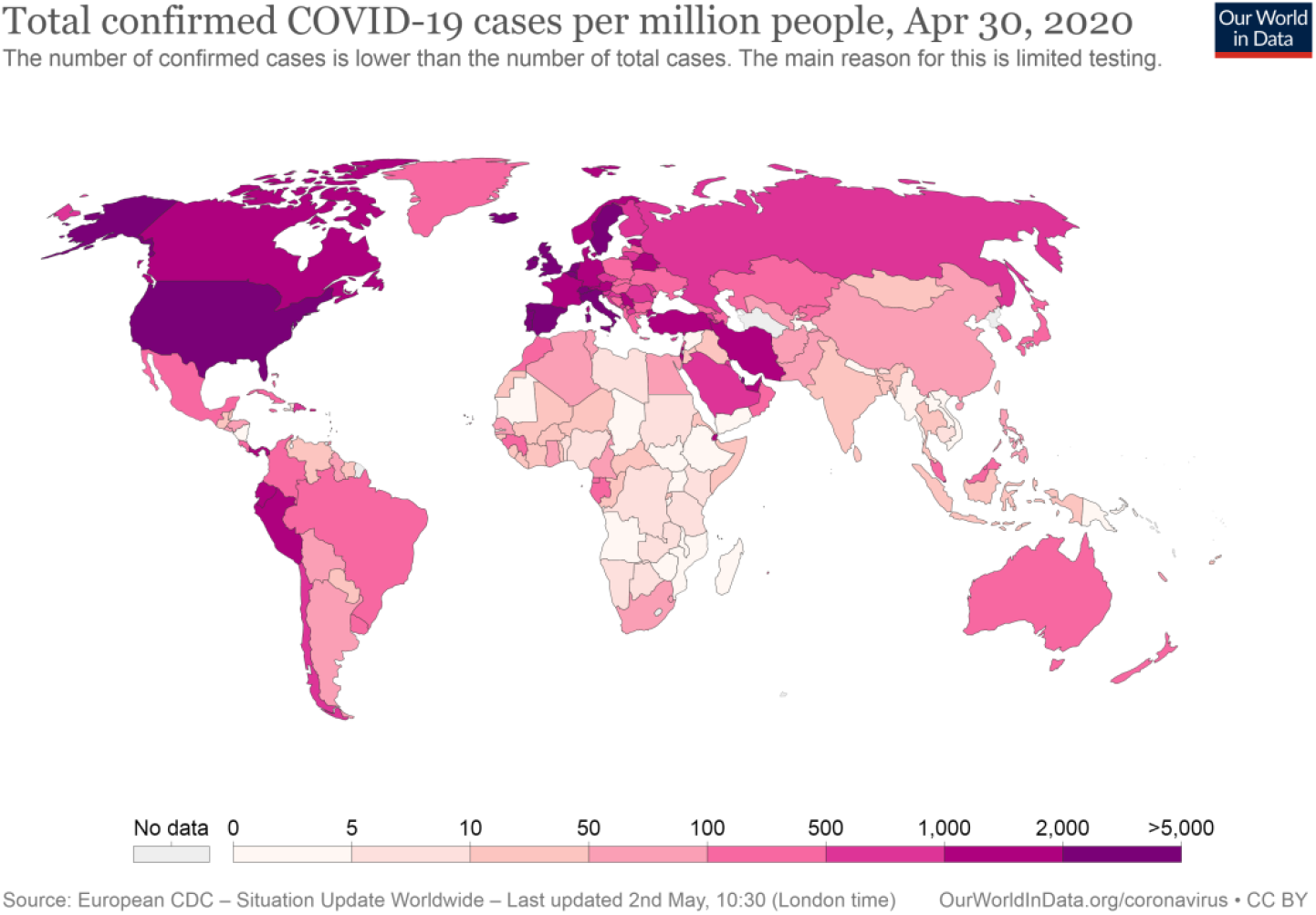
Table 3 countries with total confirmed cases of COVID-19 per million people, 30 April, 2020. Source: Our World in Data.

Considering the nonlinear aspects and variables of COVID-19 transmission and prevention of all factors such as health infrastructure facilities, new design of workflows/structures to prevent infection in health facilities, PEE (Personal protective equipment) type and availability, public health policies adopted by country, population genetics, COVID-19 testing availability and rapid response, social isolation, social distancing, economic wave influencing some essential and non-essential services/products production, government policies for supporting population and survivability, citizen collaboration to social isolation, and other public health/other policies; The author did not aim to produce statistical numerical results due to the likely lack of significance of data correlation (heteroskedasticity) for the proof that results are due to only policy interventions. All those nonlinear aspects mentioned affect the epidemics in different ways and if considering three bullet points that is: how much time does the infection occurred, which was the maximum infected population range and how many people per million; these questions addresses specifically preventive measures and for this topic, policy can be considered as the main countermeasure to keep this question answers at low standards. Therefore, it is likely that no statistical analysis with numerical results will provide any important information about community transmission among countries due to the time period limitations of the data, but mainly the nonlinear properties of the variables described necessary to predict daily new virus cases in each country. For this reason, an approach to policies influence on daily new cases was roughly described by filtering other factors that do not present high potential to accommodate the nonlinear scenario of the disease. In this sense this research results pointed that policies affected directly the population and also it can influence many of these nonlinear set of variables described before (convergence aspect of the high order non autonomous functions).

However, an overview was provided for the non-parametrical data to denote policies strong influence at the overall scenario. Note that besides this research did not focus on statistical numerical results for all set of variables of the phenomenon, these inferences were done in terms of the conceptualization of *z*- and *p*-value tests, standard deviance, variance analysis, and linear regression analysis for the policies among selected countries as it will be demonstrated at Section 4. Discussion.

The COVID-19 event was considered from a theoretical viewpoint using the qualitative theory of differential equations (QED) framework to help to understand how the input of many variables and output results in terms of convergence and stability aspects of policies adopted by each country, could point to any possible data related to empirical evidences of countries divergences visible through daily new cases and maximum time for an exponential behavior of infection. However results presented can identify true differences in terms of policies adopted and the parameters mentioned before, future studies from this point of view are necessary for other filtering aspects.

## 4. Discussion

Note that digital behavior (infodemics) [58] was not considered here, despite its potentially highly influential effect on virus transmission due to fake information and misuse of scientific information. This is a limitation of this research since even if a country has adopted all the necessary measures, citizens can undermine it. But this should be considered case by case and it does not prove the results findings directly. Future research will be conducted to integrate infodemics and life course epidemiology with a mathematical overview of the SARS-CoV-2 pandemic. This is an important tool for policy making to prevent fake news and misuse of scientific information within human behavior at the individual and community scales [58,59].

These statements lead also to the evidences, reported by the number of infected at John Hopkins University Coronavirus Resource Centre [60], that not only 50 thousand, but even 20 infected individual hosts is at risk of keeping propagating the disease [42] as observed by end of March, in China and Japan, 2020. Following this recurrence pattern, another study limitation is the time at which countries were analyzed. A longer time period is essential for obtain more accurate data for the majority of variables described in Section 3. Nevertheless, statistical data presented in this research have a strong indication of social distancing fails within some countries and success for others with the additional preventive framework of masks and city disinfection.

The observed asymptotic instability aspect of the statistical data of Figures 5-8, presenting lower infection rates for some countries (China and South Korea) while exponential infection rate for other countries, was inferred as the virus asymptote behavior to the emergent phenomenon [30-34,61,62] of community infection based on social distancing fails and the host’s social transmission behavior based or not on the use of masks and city disinfection, hence, being this theoretical mathematical model observation directly affecting the proportion of the virus reproductive formed pattern in the figure 1 and 2 listed countries.

Also, to conclude the definition of nonlinearity of SARS-CoV-2 transmission, the human emergent social transmission behavior phenomenon shares with the SARS-CoV-2, the intrinsic relation between the nonlinear expression of atmosphere possible turbulences patterns of transmission plus human random forms of socializing (subjectivity). It can results into a high convergence of infection with results pointing in the direction and the view that human most basic form of socializing, even if physically distanced, is also a tool for the self-spreading pattern of transmission [9-16,30-37,51,53,54,62,63] being this virus reproductive behavior pattern the own human social behavior itself in a very closed interaction [27-29].

This natural phenomenon of extreme convergence (host emergent social transmission behavior, virulence properties, and pathogen reproductive patterns formation) provides insight into biological organizations, and the rare extreme conditions where nature and/or human society can share uncontrolled biological association of rapid dissemination. This observation provides information about the humanity capacity to organize and to prevent and determine extreme convergence phenomena with rapid preventive and corrective responses to new high convergent pathogens [43]. The main research results are depicted in Figure 10.

**Figure 10.**
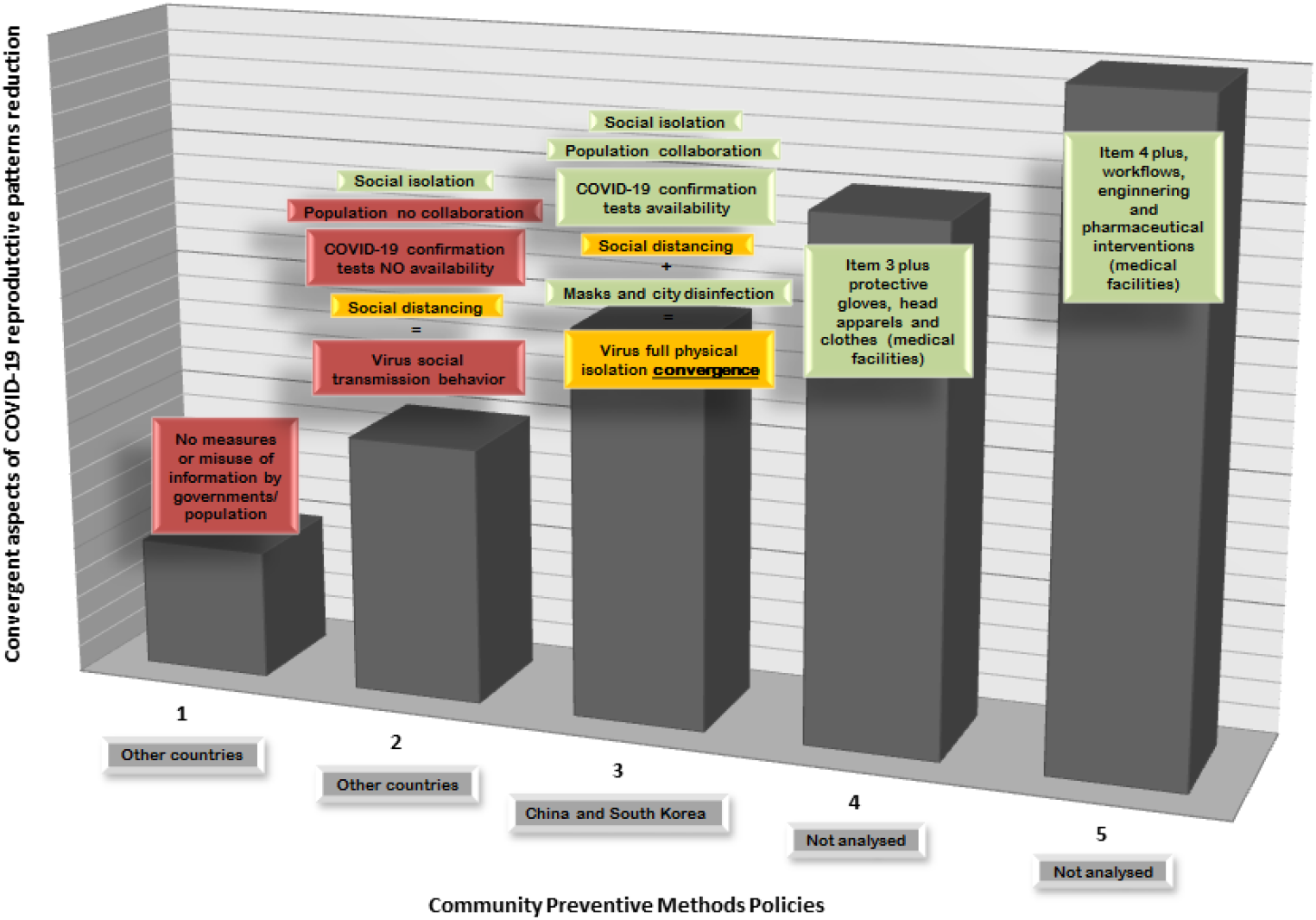
General framework of preventive methods adopted by countries and its rank of effectiveness according to data presented in table 2 and 3 results.

Item 2 in Figure 10 shows that even with social distancing measures, social transmission isolation does not occur due to the physical contact that still occurs due to atmospheric proximity. In this sense, there is no social transmission isolation of the virus spreading patterns through the atmosphere. In the next item 3, social transmission isolation is conferred by atmospheric barriers, hence, becoming the physical full isolation, even if with physical proximity, very effective.

While this research was being conducted, the daily new cases in European countries started declining (31 March, 2020). This can be attributed to the effect of social isolation policy and better results can be expected in the following days. However, this research results points to the China and South Korea different measures experience. The maximum range of infection reduction by social isolation is limited since many workforce sectors are still active. As an additional contribution to the minimum rate of virus spreading patterns that might be observed in future days, this research recommend that active citizens make use of masks [63] and countries should start to disinfect public spaces including mass transport and routes. These measures will require the introduction of policies to relax the lockdown cities by strategically and gradually allowing the population outside their homes with additional new social distancing preventive methods. Also it is necessary to observe that the minimum possible rate of new infections occurring after a city successfully reached very low rate of virus spreading patterns have the theoretical potential even under the <00,5 statistical significance of epidemics restarting (figure 11) as it was described in the last paragraph of section 3. Results, and these outcomes can be exactly in the social distancing failure approach without additional methods. But, very important to say, not uniquely caused by it.

**Figure 11.**
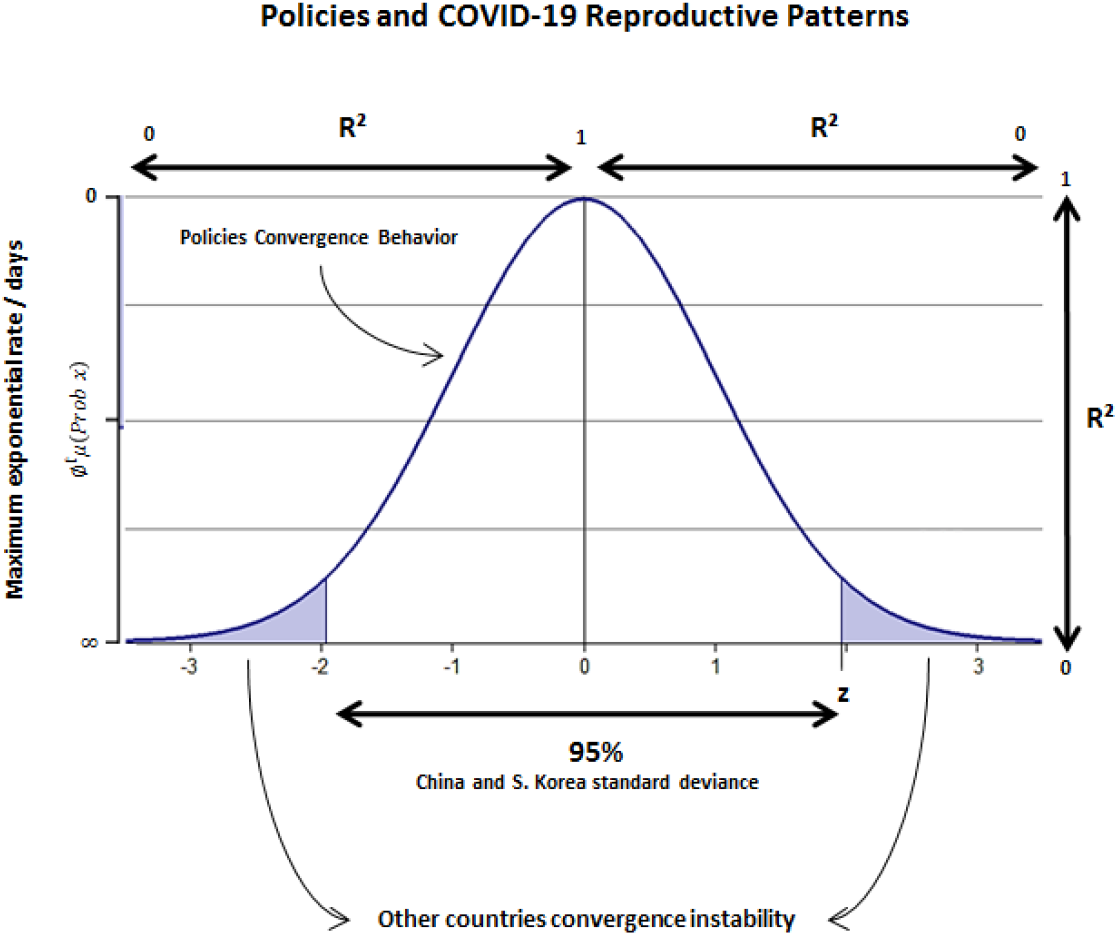
Representative scheme of SARS-CoV-2 reproductive patterns among countries whose policies might or not converge towards very low maximum exponential rate of infection per population/days. Note that countries with low max. exp. rate (table 2) also present active infection patterns, being this feature a non convergence of policies adopted, hence, expressing an exponential probability of infection constant growth (false null hypothesis).

In Figure 11, representatively, the nonlinear behavior of COVID-19 in preventive policies methodologies is represented as mandatory measures to be adopted. And also despite policies also can not grant full preventive barrier for the virus spreading in minimum rate, it can give by the experience of China and South Korea great confidence of how to keep virus exponential growth with low rate.

## 5. Conclusion

In this study, countries COVID-19 preventive policy measures were theoretically and empirically investigated; the results showed that virus reproductive patterns are tightly linked with the human social behavior and therefore, preventive policies measures and individual behavior.

Countries that adopted policy measures based on COVID-19 evidences of atmospheric potential to contaminate presented the following results:

- lower local epidemic duration;
- lower cases per 1 million;
- lower maximum exponential growth mean rate per population;
- and higher failure rate of the COVID-19 daily new cases over time.

Concerning policy measures as a whole action, social isolation and COVID-19 testing availability are mandatory for any country policy since they are the most reliable and convergent forms of obtaining best desirable solutions to reduce community virus transmission and flatten the curve goals. Concerning transmission isolation observed in China and South Korea and the superspreading patterns observed in other countries, the research identifies as full convergence of nonlinear variables for higher virus infection reduction affecting the input-output of SARS-CoV-2 propagation over time with the adoption of social isolation, COVID-19 testing availability and the social distancing methods of 1 to 2 meters physical distance with additional use of masks use and sanitization (city disinfection). Remarkably, China and South Korea presented this policy measure in contrast to other countries since beginning of epidemics and it conferred to these countries better results in controlling the local epidemics.

This reducing number of virus propagation has as a counterproof for China preventive measure, the results observed for South Korea. Other countries that didn’t follow this procedure presented high nonlinear outputs of SARS-CoV-2 transmission, mainly presenting as a common characteristic among them, the presence of constant infection cases day by day with the use of social distancing measures and with no use of all citizens masks and city disinfection. This observation leads to the hypothesis of virus atmospheric transmission potential beyond the given social distancing measures of 1 or 2 meters long, something that was also proved to be truth recently by the research of Liu et al [14] on 27 April, 2020.

Also, other important point for masks use if that if COVID-19 testing are not fully available, social distancing measures with the use of masks and city disinfection help prevent increasing daily cases, since they help to isolate the undetected infected hosts (also asymptomatic cases) and prevent airborne transmission as well as protect the uninfected hosts from environmental transmission.

This means that policy actions need to include strategic measures to address community transmission to achieve higher efficiency if social distancing measures are being maintained, including gradually the use of masks and public space disinfection measures. The lack of masks stock and financial costs, depending on the desirable level of mass masking action/disinfection, might be limiting factors. The individual dimension of prevention (citizen collaboration and support) is necessary for government actions to result in community prevention stability.

While this research was being conducted on March, some European countries analyzed in this study implemented city disinfection and mask use policies, which will likely help to reduce atmosphere and ground impact of SARS-CoV-2 transmission while, notably, in Brazil’s social isolation policies, where it will strongly impact infection reduction, is being ignored by some citizens and country president public actions.

## Data Availability

All data used in the manuscript is along the text.

## Supplementary Materials

All data are available at Statista, Our World in Data and Worldometer websites.

## Author Contributions

Not applicable.

## Acknowledgments

I would like to thank Marilei Santos da Silva, Head of Finance Department and Secretary of State for Education and Sport of Paraná for helping to disseminate research within the institution. I would like to thank all journals that have made COVID-19 research free to read, responding the call to action from the Office of Science and Technology Policy (OSTP) and other governments. This policy action was vital for researchers around the world to publish new discoveries about the virus.

